# CoSIR: Optimal control of SIR epidemic dynamics by mapping to Lotka-Volterra System

**DOI:** 10.1101/2020.11.10.20211995

**Authors:** Harsh Maheshwari, Shreyas Shetty, Nayana Bannur, Srujana Merugu

**Affiliations:** Department of Computer Science, Georgia Insitute of Technology; Flipkart Internet Pvt. Ltd.; Wadhwani AI; Independent

**Author notes:** Equal Contribution (Harsh Maheshwari, Shreyas Shetty).

## Abstract

Multiple macro-phenomena such as disease epidemics, online information propagation, and economic activity can be well-approximated using simple dynamical systems. Shaping these phenomena with adaptive control of key levers has long been the holy grail of policymakers. In this paper, we focus on optimal control of transmission rate in epidemic systems following the widely applicable SIR dynamics. We first demonstrate that the SIR model with infectious patients and susceptible contacts (i.e., product of transmission rate and susceptible population) interpreted as predators and prey respectively reduces to a Lotka-Volterra (LV) predator-prey model. The modified SIR system (LVSIR) has a stable equilibrium point, an “energy” conservation property, and exhibits bounded cyclic behavior. We exploit this mapping using a control-Lyapunov approach to design a novel adaptive control policy (CoSIR) that nudges the SIR model to the desired equilibrium. Combining CoSIR policy with data-driven estimation of parameters and adjustments for discrete transmission levels yields a control strategy with practical utility. Empirical comparison with periodic lockdowns on simulated and real COVID-19 data demonstrates the efficacy and adaptability of our approach.

## 1 Introduction

The COVID-19 epidemic, proliferation of fake news, and increasing economic inequality all highlight the necessity and the enormous challenges of controlling social macro systems. Often, these systems can be characterized by simple dynamics parameterized along certain factors. Optimal adaptive control of such factors is a critical problem for policy makers with massive societal consequences.

Management of a pandemic is a prime example of such a control problem with four key levers: (a) contact restrictions, (b) testing, tracing and isolation, (c) provisioning for medical capacity, and (d) vaccinations. Of these, contact restrictions is the most powerful policy instrument, especially in low and middle income countries facing significant resource constraints. However, choosing the optimal restrictions is highly non-trivial not only because of the complex trade-off between the disease impact and socioeconomic disruptions but also due to the rapidly evolving situation on the ground.

Public health interventions related to COVID-19 have largely been driven by scenario-based epidemiological forecasting [Ferguson et al., 2020, Ray et al., 2020] with decision-making based on comparison of a limited number of scenarios over a finite time horizon. Though valuable, this approach leans towards a reactive role for the health authorities. In contrast, despite the potentially far-reaching impact, relatively less attention has been devoted to developing an analytic control framework to support proactive decision-making based on the target disease and economic outcomes, and the state of the epidemic. Multiple studies [Chowdhury et al., 2020, Killian et al., 2020, Bin et al., 2021] point to the benefits of periodic lockdowns, but these interventions are based on simulations over restricted scenarios and are not adaptive in nature. Some recent works [Acemoglu et al., 2020] formulate the control problem in terms of net socioeconomic and disease impact but require an expensive numerical solution. Recent reinforcement learning-based formulations [Bhardwaj et al., 2020] are efforts in similar direction but do not fully exploit the mathematical structure of the epidemic dynamics.

In this paper, we explore optimal adaptive control of transmission rate for a desired bound on infectious population. Multiple studies [Chen et al., 2020, Ray et al., 2020] indicate that time-varying SIR and SEIR [Hethcote, 2000] compartmental models permit parsimonious encoding of infection dynamics, data-driven calibration, and accurate forecasts leading to wide adoption of these models. Hence, we focus on the SIR model [Kermack and McKendrick, 1927] for the control analysis.

### Contributions

1. [**Section 4**] We show that the SIR dynamics maps to the well-known Lotka-Volterra (LV) system [Baigent, 2016] on interpreting infectious patients as predators and susceptible contacts (the product of transmission rate and susceptible population) as the prey under a specific transmission rate policy. The resulting system (LVSIR) has a stable equilibrium point and an “energy” conservation property. It exhibits a bounded cyclic trend for active infections and a steady decline of susceptibility.
2. [**Section 5**] We exploit the mapping to derive optimal control policy for transmission rate (`CoSIR`) using control-Lyapunov functions [Tsinias, 1988] based on the “LV system energy”. The policy is guaranteed to converge to the desired equilibrium (target infection level) from any valid initial state assuming stochastic perturbations. ^2^
3. [**Section 6**] We propose a hybrid practical strategy that combines `CoSIR` with data-driven estimation of parameters and restriction to discrete levels (`CoSIR-approx`).
4. [**Section 7**] Empirical results on real and synthetic COVID-19 data demonstrate the efficacy of the CoSIR approach in stabilizing infections and adaptability to perturbations.

Table 1 lists the relevant SIR models for clarity. Our work points to a general control strategy: (a) mapping a real-world phenomenon to a well-behaved Hamiltonian system [Nutku, 1990] amenable for control, (b) designing optimal control mechanisms for levers of interest using control theory, and (c) employing learning methods to estimate the other model parameters from data. Other application domains such as online information propagation could benefit from a similar hybrid approach.

**Table 1:** Variants of the relevant SIR models and transmission rate *β*.

|  |  |
| --- | --- |
| SIR (Constant $\beta$) | $\beta$ is constant |
| Lotka Volterra-SIR (LVSIR) | $\beta$ varies with time following Eqn. 2 |
| Controlled SIR (CoSIR) | $\beta$ varies with time following control mechanism in Eqn. 3 |
| Approximate Control (CoSIR-approx) | $\beta$ from CoSIR is approximated by $k$ discrete levels |

**Notation**: *x*_*t*_ and *x*(*t*) both denote the value of *x* at time *t*; 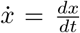 denotes the time derivative; [*x*_*t*_], 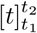 denotes the series from *t* = *t*_1_ to *t* = *t*_2_.

## 2 Problem Formulation

During an epidemic, a key concern for public health officials is to determine the right level and schedule of contact restrictions that balance the disease and socioeconomic burdens. Strict short-term lockdowns suppress the infection levels but infections tend to rise on easing restrictions unless the epidemic is completely wiped out. On the other hand, prolonged restrictions with no intermittent easing hinder economic activity and impose heavy costs on vulnerable population groups.

Modeling the multi-faceted impact of contact restrictions requires accounting for region-specific cultural and economic constructs as well as the available resources, which is highly complex. For tractability, we assume that the public health goal is to limit active infections to a certain target level determined via an independent impact analysis [BMGF, 2020]. The controls available to the public health authorities can be viewed as configurable knobs (e.g., offices at 20% occupancy). However, the need for clear communication and public compliance entails a simpler strategy centered around a few discrete restriction levels (see Table 6), and a preset schedule for a future time horizon.

### Restriction Policy Optimization

For a given region, let (*N, S*_*curr*_, *I*_*curr*_) be the total, current susceptible, and infectious populations respectively. Let 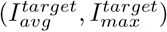 be the target average and maximum infectious levels. Let 𝒜 be the set of restriction levels for which the transmission rate is known or can be estimated^3^ and *T*, the decision horizon. The goal is to identify restriction levels [*a*_*t*_], 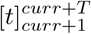, *a*_*t*_ ∈ 𝒜 such that the infectious level averages at 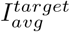 but does not exceed 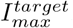. In a general case, the target levels could themselves be time-varying functions instead of static values.

Since our goal is to develop an analytical control framework, we make simplifying assumptions on the observability, (i.e., accurate estimation of the infectious population via sero-surveys and diagnostic tests) and the disease dynamics (homogeneous interactions, negligible incubation, constant infectious period). Appendix A describes extensions when some of these assumptions are relaxed.

## 3 Preliminaries

### Compartmental Models

Infectious diseases are often modeled using compartmental models where the population (*N*) is divided into compartments corresponding to disease stages with transitions governed by the model dynamics. The SIR model [Kermack and McKendrick, 1927] is the simplest and most widely used one comprising three compartments: Susceptible (**S**), Infectious (**I**) and Removed (**R** - includes immune &post-infectious persons) with the dynamics in Figure 1(b). Here *β* is the rate of disease transmission from infectious to susceptible individuals, which largely depends on the contact restrictions and *γ* is the inverse of the average infectious period. The effective reproduction number (average number of direct infections from each infection) is 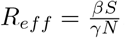. Existing restriction control approaches [Systrom et al., 2020] are often guided by the principle of ensuring *R*_*eff*_ ≃ 1.

**Figure 1:**
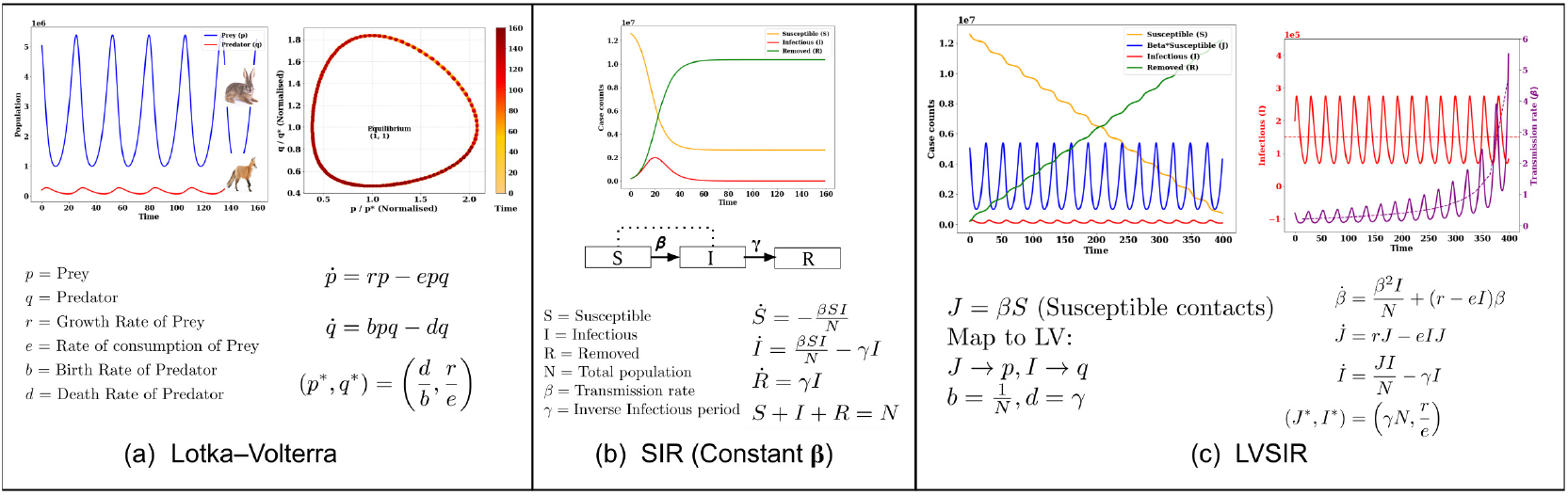
(a) LV system and time evolution of predator and prey populations, (b) SIR dynamics with constant *β*, (c) SIR model to LV system mapping and the behaviour of case counts *S, J, I, R* and transmission rate *β*.^4^

### Lotka-Volterra (LV) Systems

LV systems [Baigent, 2016] model the population dynamics of predator-prey interactions in an ecosystem. In a simple two-species LV system, the population of prey (*p*) interacts with that of predator (*q*). The growth rate of prey depends on its reproduction rate (*r*) ^5^ and the rate of consumption by predator (*e*). The change in predator population depends on the nourishment-based birth rate *b* and its death rate *d*. The system has two fixed points: (a) a saddle point that maps to extinction (*p, q*) = (0, 0), and (b) a stable equilibrium at 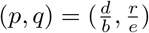. Typically, the system exhibits oscillations resulting in a closed phase plot that corresponds to conservation of an “energy” function. Figure 1(a) presents the dynamics of an LV system and the oscillations of the prey and predator populations. Due to the criticality of ecological population control, there has been considerable research on multiple variants of LV systems [Hening and Nguyen, 2018, Wangersky and Cunningham, 1957] and their Hamiltonian dynamics [Nutku, 1990].

### Optimal Control Strategies

Optimal control of dynamical systems Powell [2019] is a well-studied area with connections to reinforcement learning. Given a set of control variables, the optimal control policy describes the time derivatives that minimize the cost function and can be derived using Pontryagin’s maximum principle [Powell, 2019] or the Hamilton-Jacobi-Bellman equations [Peng, 1992]. In the case of linear dynamical systems, there are straightforward techniques for optimal control. However, control of non-linear dynamical systems relies heavily on the existence of control-Lyapunov functions often identified using conservation laws of the associated systems, and is non-trivial. Once a suitable Lyapunov function is identified, there exist multiple control design methods such as feedback linearization, backstepping and sliding mode control that are guaranteed to converge using Artstein’s theorem [Artstein, 1983]. In the case of the SIR model, a suitable Lyapunov function is not readily evident. On the other hand, Lyapunov stability and practical control strategies of LV systems have been extensively studied [Meza et al., 2005, Pchelkina and Fradkov, 2012].

## 4 Mapping SIR to Lotka-Volterra System (LVSIR)

Solving the restriction policy optimization is our primary goal. We focus on control of SIR dynamics as it captures the core disease spread mechanism of most epidemiological models.

Since existing work [Adda and Bichara, 2012] addresses controllability of SIR dynamics only in the limited context of endemic equilibrium, we adopt a new approach. We first establish a connection between the SIR dynamics and Lotka-Volterra (LV) system and then leverage the LV system properties to propose a strategy for transmission rate control in the SIR system (Section 5).

Stabilizing infection levels has a direct analogy with population control in LV predator-prey systems where it is desirable to maintain the predator and prey population at certain target levels. Comparing the SIR and LV dynamics in Figure 1, we observe that the behaviour of the infectious people (*I*) is similar to that of the “predators” (*q*). There is an inflow (birth) *βSI/N* that depends on *β* as well as the current infectious and susceptible population. There is also an outflow (death) *γI* from the **I** to the **R** compartment. However, the counterpart for the “prey” is not readily apparent.

An intuitive choice for “prey” is the “susceptible contacts” (i.e., the product of susceptible people and *β*, the number of “contacts” of a person per day) since this acts as “nourishment” to the predators and contributes to the inflow into the **I** compartment. Denoting the susceptible contacts by *J* := *βS*, we note that equivalence with the LV system requires

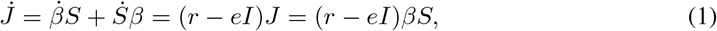

where *r* and *e* correspond to the reproduction rate and consumption rate of an LV system as described in Section 3.2. Since 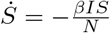, we require the transmission rate *β* to follow

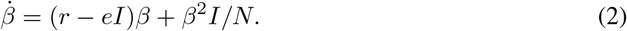

This modified version of SIR model (LVSIR) maps to an LV system. The inverse of the infectious period (*γ*) corresponds to the predator death rate (*d*) and the inverse of population (1*/N*) to the birth rate (*b*). The LV reproductive rate (*r*) and prey consumption rate (*e*) are extra degrees of freedom.

Theorem 1 asserts the existence of an equilibrium for the LVSIR system and the associated behavior.

### Theorem 1

^2^ *For the LVSIR model in Figure 1(c), the following holds true:*

1. *There exists a stable equilibrium at* (*J*\*, *I*\*) = (*γN, r/e*).
2. *When the initial state* (*J*_0_, *I*_0_) = (*J*\*, *I*\*), *then* (*J, I*) *remain constant while S, R take a linear form and β increases till the susceptible population reaches 0 at T*_*end*_.
  i. *S*(*t*) = *S*_0_ − *γI*\**t*; *R*(*t*) = *R*_0_ + *γI*\**t*
  ii. 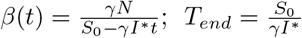

**Proof sketch**. (1) Setting time derivatives to 0 yields the equilibrium points. The stability emerges from the conservation of the “system energy” as defined in Theorem 2(a) and its strict convexity. (2) Invoking the model dynamics (Figure 1(b)) on initial state yields the result.

When the LVSIR system is initialized at a non-equilibrium state, it exhibits an oscillatory behavior and a conservation property, which are characterized in Theorem 2 below.

### Theorem 2

^2^ *For the LVSIR model in Figure 1(c), if the initial state is not at equilibrium, i*.*e*., (*J*_0_, *I*_0_) ≠ (*J*\*, *I*\*), *it exhibits a cyclic behaviour with the following properties*.

1. *The system “energy” w*(*J, I*) = *γ*(*x* −log(*x*) − 1) + *r*(*y* −log(*y*) − 1) *(where* 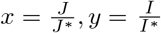 *) remains constant at w* = *w*(*J*_0_, *I*_0_) ≥ *w*(*J*\*, *I*\*) = *w*\* = 0 *till termination*.
2. *The I, J curves exhibit periodic oscillations resulting in a closed trajectory. The normalized phase plot has four extreme points* {(*x*_*min*_, 1), (1, *y*_*min*_), (*x*_*max*_, 1), (1, *y*_*max*_)} *where* (*x*_*min*_, *x*_*max*_) *are the roots of the equation x* − log(*x*) = 1 + *w*_0_*/γ and* (*y*_*min*_, *y*_*max*_) *are the roots of the equation y* − log(*y*) = 1 + *w*_0_*/r*.
3. *The cyclic period is given by* 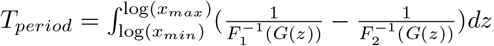, *where* (*x*_*min*_, *x*_*max*_) *are defined as above, G*(*z*) = *γ*(*e*^*z*^ −*z* −1)−*w*_0_, *and F*_1_(*s*), *F*_2_(*s*) *are restrictions of* 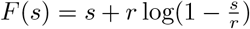 *to positive and negative ranges. In general, T*_*period*_ *has the form* 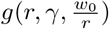 *with approximation via linearization yielding* 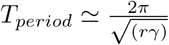.
4. *In each cyclic period, S reduces by a fixed amount* Δ*S* = *γI*\**T*_*period*_. *When S* < Δ*S at the start of a cycle, the epidemic terminates during that cycle*.

**Proof sketch**. (1) Substituting the model dynamics into the energy function yields time-invariance. (2) The closed phase plot emerges from the conservation property with the extreme points determined by the properties of the convex function *f* (*x*) = *x* − log(*x*) − 1. (3) Derivation of the time period follows a similar analysis for Lotka-Volterra systems [Hsu, 1983]. (4) Fixed drop in *S* follows from the periodicity and the fact that *I*\* is the average value of *I* in each cycle.

Figure 2(a) depicts the oscillatory behaviour of the LVSIR model for the hypothetical region in Table 3. Similar to an LV system, the “energy” which corresponds to a weighted Itakura-Saito distance [Itakura and Saito, 1968] between (*I, J*) and the equilibrium (*I*\*, *J*\*) is conserved. The infectious population *I* (and susceptible contacts *J*) oscillates between [*y*_*min*_*I*\*, *y*_*max*_*I*\*] (and [*x*_*min*_*J*\*, *x*_*max*_*J*\*]) during the entire period with an average value of *I*\* (and *J*\*) while the susceptible population reduces steadily in a staircase-like fashion, which is desirable from a public health perspective. The transmission rate *β* also exhibits periodic oscillations but the average steadily goes up to compensate for the reduction in the susceptible population. Figures 2(b-c) show the phase plot and the variation of the key quantities during a single period while Figure 2(f) shows dependence of *T*_*period*_ on *r* and *w*_0_*/r*.

### Algorithm 1 Practical Transmission Rate Control

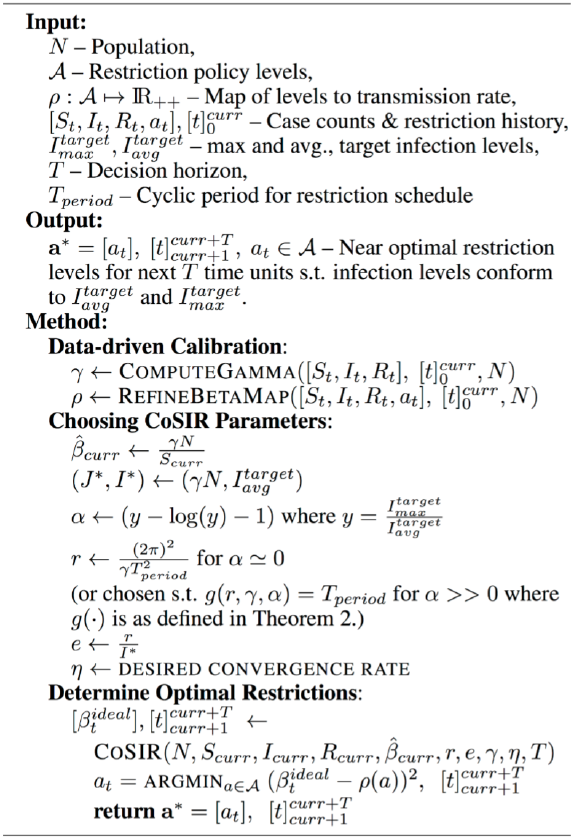

**Figure 2:**
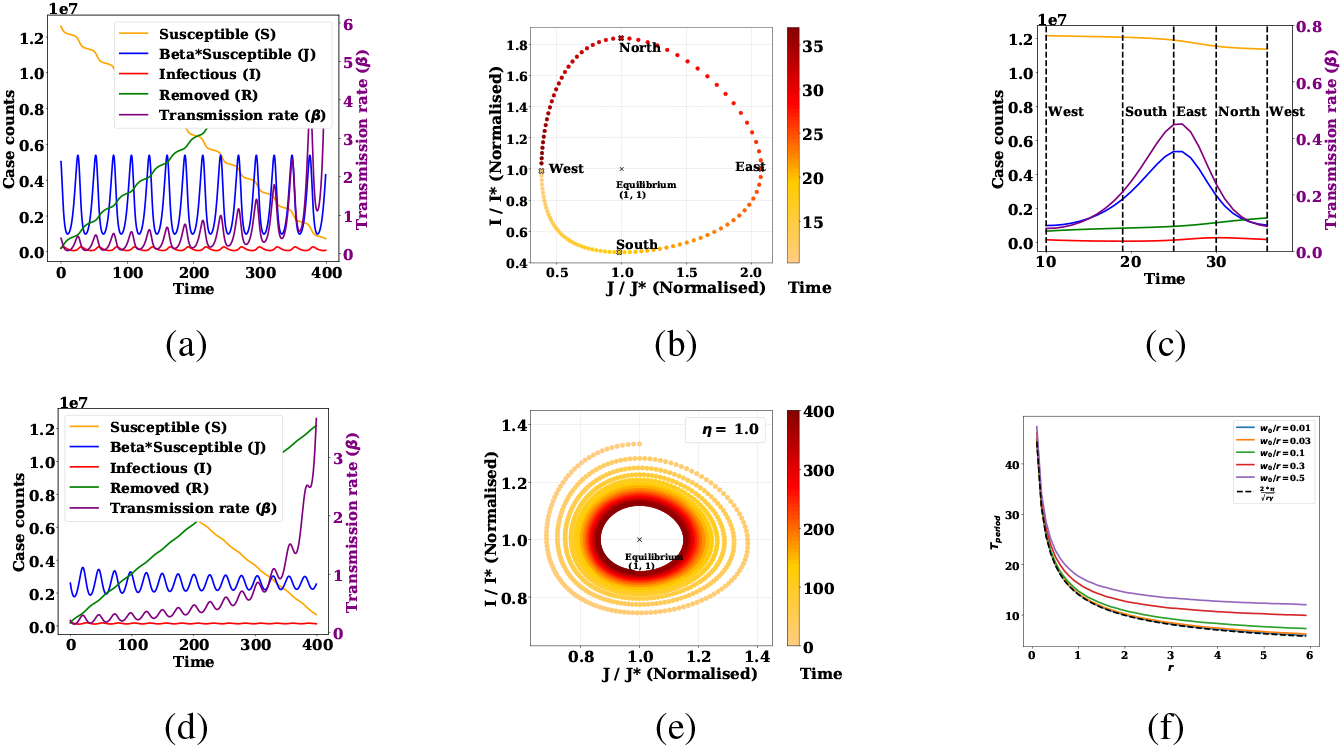
(a-c) System (*S, J, I, R, β*) evolution when *β* follows Eqn. 2 [LVSIR] with its phase plot and single cycle annotated with extreme points, (d-e) System evolution when *β* follows Eqn. 3 [CoSIR] and learning rate *η* = 1, (f) Dependence of *T*_*period*_ on *r* for different choices of *w*_0_*/r*.^2^

## 5 Transmission Rate Control (CoSIR)

Consider an epidemic system from a control theory perspective with the compartmental populations as the system state, the transmission rate as the control input, and the external flows as perturbations. The SIR system has a natural positive feedback loop because of the infection mechanism (since 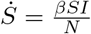) which leads to an exponential-like behavior in the early phase of the epidemic. The LVSIR dynamics neutralizes this feedback loop to create oscillatory behavior. We now explore the problem of controlling the transmission rate *β* to nudge the infectious levels to a desired equilibrium.

As discussed in Section 3, control of non-linear dynamical systems is typically achieved via Control-Lyapunov functions (CLFs) defined below. Hence, our immediate goal is to exploit the SIR-LV mapping to derive CLFs based on the “Lotka-Volterra energy” function *w*(*J, I*) in Theorem 2.

### Definition 1

*Given a dynamical system* **ż** = **f**(**z, u**) *with state vector* **z** ∈ *D* ⊂ ℝ^n^, *control* **u** ∈ ℝ^m^, *and equilibrium state* **z**\* = **0**, *a control-Lyapunov function (CLF) is a function V* : *D* ↦ ℝ *that is continuously differentiable, positive-definite s*.*t*.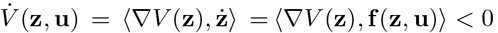.

The CLF *V* (·) can be viewed as a generalized energy function with 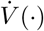 being a dissipation function. Artstein [1983] proved that as long as there is a CLF, there exists a control **u** to ensure the reduction of energy at every non-equilibrium state and eventual convergence to the zero energy equilibrium.

### Theorem 3

**(Artstein [1983])** *For any non-linear dynamical system with affine control*, 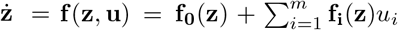 *with state* **z** ∈ *D* ⊂ ℝ^n^, *control* **u** ∈ ℝ^m^, *has a CLF iff it admits a regular stabilizing control feedback* **u**, *that is a locally Lipschitz function on* ℝ^n^\ {0}.

Once a CLF is identified, it is relatively straightforward to design an appropriate control function **u** as described in [Artstein, 1983, Tsinias, 1989]. For the current scenario, we consider appropriately defined functions of the LV energy. Let 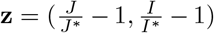 so that the equilibrium **z**\* = (0, 0) and *L*(*w*) : ℝ_+_ ↦ ℝ be a continuously differentiable function such that 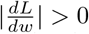 and *L*(*w*) ≥ 0, ∀*w* and *L*(*w*) = 0 ⇔ *w* = 0. Then, the function *V* (**z**) = *L*(*w*(*J, I*)) can be shown to be a CLF for the SIR model and used to construct an affine control policy (CoSIR) as described below.

### Theorem 4

**(CoSIR)** ^2^ *For the SIR model, a proportional additive control on β of the form*,

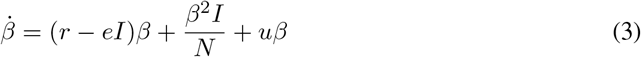

*converges to the equilibrium* (*J*\*, *I*\*) *when* 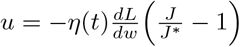 *with learning rate η*(*t*) > 0, ∀*t*.

**Proof sketch**. Follows from Definition 1 and Theorem 3 using 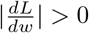.

Pchelkina and Fradkov [2012] proposed a special case of the above construction with 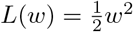, which is referred to as the speed-gradient method. Figures 2(d-e) show the damping oscillatory behavior of the CoSIR model for this special case, for the hypothetical region in Table 3. The *β*-control policy (Eqn. 3) can be interpreted as follows. The first term 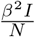 corresponds to the relaxation possible due to the decreasing susceptible population while the second term (*r* − *eI*)*β* leads to oscillatory behavior, and the last term 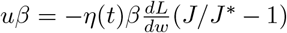 ensures dissipation of energy and convergence to the equilibrium. Even with external perturbations, the system can recalibrate *β*, but the convergence properties depend on the nature of perturbations and need to be explored.

## 6 Practical Transmission Rate Control

Algorithm 1 outlines a practical approach to solve the restriction policy optimization problem (Section 2) using the optimal *β*-control in Theorem 4. There are four key steps.

### Input Collection

Infection level targets 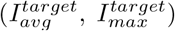, periodicity of the restriction schedule (*T*_*period*_), decision horizon (*T*) need to be determined based on public health and socioeconomic considerations. Historical case counts and restrictions 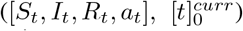 also need to be collected to enable accurate calibration and policy optimization.

### Data-driven Calibration

We then employ SIR calibration methods to estimate the parameter *γ* (ComputeGamma) and the state of the epidemic (*S*_*curr*_, *I*_*curr*_ *R*_*curr*_) from historical data. There is a huge body of literature in this space [Ray et al., 2020, Bettencourt and Ribeiro, 2008, Baek et al., 2020] based on likelihood maximization, Bayesian optimization and MCMC methods that can be readily employed. Similarly, the restriction level to transmission map (*ρ*) can be initialized from public health guidelines [COVID-AMP, 2020] and refined using the observed *β* for past restrictions (RefineBetaMap) via online learning or contextual bandit methods [Beygelzimer et al., 2011]. For the sake of brevity, we skip a detailed description of these techniques (more details in Appendix D).

### Choosing COSIR Parameters

The free parameters of the COSIR model need to be chosen based on the control requirements. Algorithm 1 lists the choices derived from Theorem 2. Equilibrium *I*\* is chosen to be the same as the average target infectious level, LV reproductive rate (*r*) is based on the desired cyclic period *T*_*period*_ and the ratio of the maximum and average target levels, and lastly the consumption rate (*e*) is determined by *r* and *I*\*. There is flexibility on the choice of 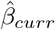 and *η*. Choosing the immediate transmission rate to be 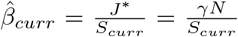 (equivalent to forcing effective reproduction number *R*_*eff*_ = 1) ensures a maximal reduction in the system “energy” and faster convergence to the desired equilibrium, but dampens fluctuations. However, fluctuations might be necessary for economic activity. When nearly steady infection levels are desired, 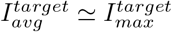, then 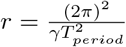 and high *η* are appropriate.

### Computing Near Optimal Restrictions

Determining the restriction policy can be split into two phases. The first involves estimating the ideal *β* control (`CoSIR`) from Eqn. 3 while the second involves identifying the “closest” restriction level for the ideal *β* at each time step with “closeness” based on a suitable divergence such as the squared loss (`CoSIR-approx`).

The main utility of the above approach is that it combines estimation of model parameters (ComputeGamma, RefineBetaMap) with the underlying system dynamics to arrive at a practical strategy. Additional aspects relevant for a real-world implementation are discussed in Section 9.

## 7 Empirical Results

We present simulation results on the COVID-19 pandemic to demonstrate the efficacy and adaptability of the proposed `CoSIR` approaches relative to other widely cited approaches [Chowdhury et al., 2020].

### Experimental Setup

#### Data

Simulated data (Apr 2020-Apr 2021) from two regions (India, Netherlands) was used along with real COVID-19 data [Dong et al., 2020] from the initial phase of pandemic (till end of Dec 2020) where the SIR dynamics are more applicable due to negligible reinfection and vaccination rates.

#### Algorithms

We consider five transmission control policies (see Table 2) assuming SIR dynamics. These include a completely non-restrictive policy (`No-Restrictions`), two CoSIR-based policies (`CoSIR`, `CoSIR-approx`), and two baseline policies (`PL-low`, `PL-high`) based on dynamic periodic interventions [Chowdhury et al., 2020]. We also include the real outcome as indicative of the actual public health transmission control that was adopted.

**Table 2:** Transmission control policies used for simulation.

| Algorithm | Description |
| --- | --- |
| No-Restrictions | Constant $\beta = 0.44$ |
| PL-high | Periodic Lockdown of 60 days with $\beta = 0.16$ and a relaxation period of 30 days with $\beta = 0.44$ |
| PL-low | Periodic Lockdown of 60 days with $\beta = 0.1$ and a relaxation period of 30 days with $\beta = 0.44$ |
| CoSIR | Follows Eqn. 3 |
| CoSIR-approx | Approximation of CoSIR $\beta$ with 10 levels starting from 0.1 to 0.55 in steps of 0.05. |

#### Parameter Choices

Table 3 enumerates the parameters chosen based on COVID-19 characteristics and practical considerations (details in Appendix C). Two critical choices in our simulations are the restriction periodicity which was set to a weekly cadence and the target infectious level 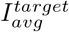 which was chosen based purely on medical capacity constraints due to the absence of other information. We estimate a manageable hospitalization inflow 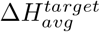 based on factors such as population and per-capita bed capacity and set 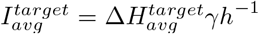 noting that in the case of SIR dynamics, the new hospitalizations equal *γhI* where *h* denotes the fraction of cases requiring hospitalization.

**Table 3:** State of the epidemic and public health requirements used for the simulations in Section 7 for a hypothetical region (HYP), India (IN) and Netherlands (NL).

| Factor | Symbol | HYP | IN | NL |
| --- | --- | --- | --- | --- |
| Population | $N$ | 13M | 1.36B | 17.1M |
| Susceptible Population | $S_{curr}$ | 12.6M | 1.36B | 17.09M |
| Infectious Population | $I_{curr}$ | 0.2M | 279k | 6.7k |
| Hospitalization ratio | $h$ | - | 2% | 6.6% |
| Target manageable I | $I_{avg}^{target}$ | 0.15M | 6.8M | 0.16M |
| Target max to avg ratio | $\frac{I_{max}^{target}}{I_{avg}^{target}}$ | 1.3 | 1.13 | 1.13 |
| Target H inflow | $\Delta H_{avg}^{target}$ | - | 27.5k | 2.1k |
| Periodicity | $T_{period}$ | 7 days | 7 days | 7 days |
| Inverse Infectious period | $\gamma$ | 0.2 | 0.2 | 0.2 |
| LV Reproduction rate | $r$ | 4.2 | 4.2 | 4.2 |
| Consumption rate | $e$ | $r/I^*$ | $r/I^*$ | $r/I^*$ |
| Learning rate | $\eta$ | 5 | 5 | 5 |

### Hospitalization Influx

Figures 3(a,d) show the simulated daily new hospitalizations (*γhI*) while Figures 3(b,e) depict the average excess and under hospital inflow relative to the target rate as follows: 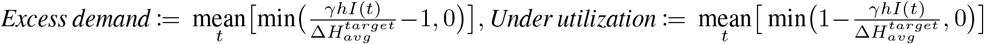. Real hospitalizations are based on appropriate scaling of the reported active cases accounting for under-reporting (Appendix C) and point to periods where the healthcare system was overwhelmed (India) or the restrictions were more stringent than necessary (Netherlands). The `CoSIR` variants result in nearly steady hospitalization inflow with the smallest deviation from the target levels relative to all other choices including the actual implemented policy. The key takeaway is that optimizing resource utilization requires choosing *β* based on the constraints and the varying susceptibility.

**Figure 3:**
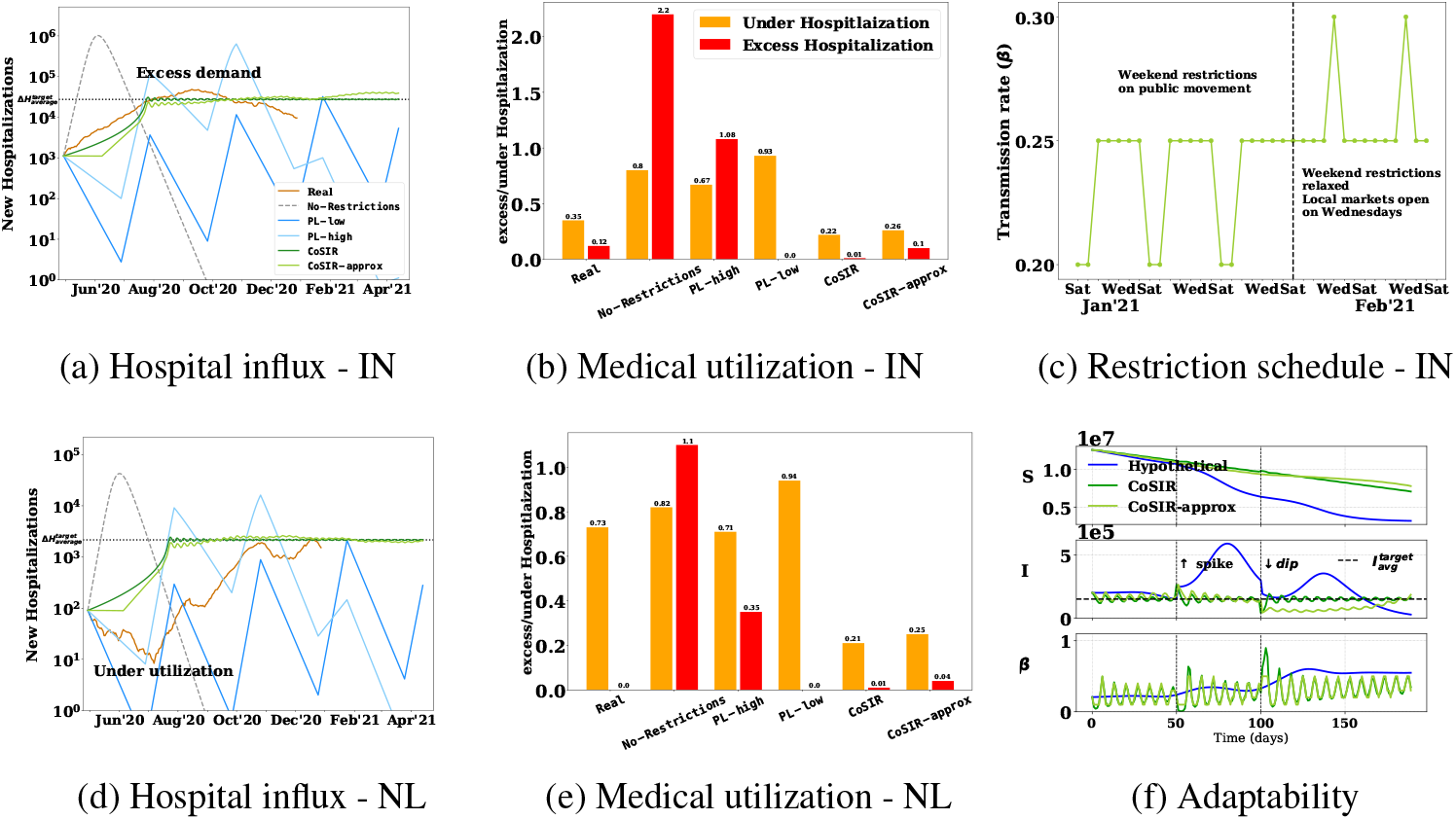
(a-b, d-e) New hospitalization influx and excess/under hospitalization with various transmission control policies for two regions: India-IN and Netherlands-NL, (c) `CoSIR-approx` schedule (IN) for a 5-week period (see Appendix F for full range) with a possible mapping to real restrictions, (f) Disease progression indicates that `CoSIR` variants adapt to target levels and also to a sudden increase (*t* = 50) or decrease (*t* = 100). ^2^

### Feasibility of CoSIR-approx Restriction Policy

Figure 3(c) shows the `CoSIR-approx` *β* schedule for a 5-week duration comprising two phases each of which has a weekly periodicity. This schedule could potentially be implemented by having tighter weekend restrictions in the initial phase which are then eased in the later phase along with permitting high contact activities (e.g., local market) on a single weekday. Public communication for these multi-week phases is quite feasible.

### Adaptability of CoSIR

Figure 3(f) shows the epidemic evolution of the two CoSIR variants along with a hypothetical but a realistic premature relaxation policy. The CoSIR variants not only stabilize infections, but are able to adapt to sudden upward (t = 50) or downward perturbations (t = 100) as in the case of super-spreader events or sudden quarantine restrictions respectively and continue pushing towards the equilibrium. In practice, the adaptation is not seamless and depends on the nature of perturbations and the lags in the data collection and public communication processes.

## 8 Related work

Most of the existing literature on epidemiological modeling focuses on the following aspects: (a) Design of models to capture the disease dynamics [Harko et al., 2014], (b) Accurate forecasting of future case counts [Ray et al., 2020], (c) Estimation of model parameters corresponding to non-pharmaceutical interventions (NPIs) [Davies et al., 2020] (d) Optimization of public-health policy (especially NPIs such as quarantine and lockdown policy) based on economic impact and disease burden [Acemoglu et al., 2020, Petrie and Masel, 2020]. There is also work [Naji and Mustafa, 2012] in the space of eco-epidemiology that interleaves SIR and LV dynamics to model prey-predator populations when there is an infection spreading among the prey but it is substantially different from the current effort. Our current work is aligned with the last area, often referred to as economic epidemiology, since the objective is identification of optimal NPIs. However, most of the research in this space is focused on the variation of disease dynamics across subgroups using compartmental models and the explicit modeling of the economic and disease impact along with the trade-offs. The formulated optimization problems are often not readily tractable. Due to the subjective nature of the socioeconomic modeling assumptions and the computationally expensive solutions, this work is more suited for static recommendations. Our work, on the other hand, focuses on dynamic adaptive control of the transmission rate assuming simple SIR dynamics with the goal of maintaining a specific (possibly varying) target level of infection. The resulting analytic control policy adjusted for practical considerations can be useful for real-time decision support.

## 9 Assumptions &Limitations

Real-world utility of the proposed control framework relies on a few key assumptions. The first pertains to the applicability of the SIR dynamics (i.e., ODEs in Figure 1 with *β* possibly varying with time) and the associated homogeneous mixing property for a given epidemic and region. This has been validated for MMR and Ebola epidemics for certain regions [Yuan et al., 2015] with SEIR model being preferable for COVID-19. The second assumption relates to the existence of a 1-1 mapping between contact restrictions and the transmission, which is somewhat validated by multiple studies [Liu et al., 2021] though the relationship can include delays and stochasticity due to compliance issues. The third assumption is on the nature of perturbations or the external flows since the optimality of control and convergence results are guaranteed only for the ideal *β* control with normal stochastic perturbations. Adaptability under arbitrary/adversarial perturbations remains to be explored. Effective implementation in a practical public health setting similar to the dynamic protocol deployed in California [CA, 2020] also depends heavily on ensuring (a) appropriateness and availability of a target (possible time-varying) infection level through an independent impact analysis, (b) robust data-driven estimation of model parameters (e.g., *γ*) as well as the restriction to *β* mapping, (c) adjusting for the lags and under-reporting in the data collection process, and (d) accounting for delays in public communication systems and varying compliance. An important limitation of our current work is that the efficacy of control strategies could only be demonstrated via generic SIR-based simulations since real experiments are impractical due to the associated human impact. As more case and restriction data becomes available, we plan to perform off-policy evaluation.

## 10 Conclusion &Future Work

We propose an analytical framework for epidemic control with the intent of supporting an active goal-oriented public health response. This framework relies on a mapping between SIR dynamics to Lotka-Volterra system under a specific transmission rate policy (LVSIR) and an additional feedback control mechanism (CoSIR). Given the vast literature on control of LV systems, this mapping can be leveraged to design new epidemic control techniques as well as extend current results to richer compartmental models and additional control variables (Appendix A). Results in diverse settings point to the utility of this approach. Qualitative insights such as weekly restriction patterns were employed in a regional COVID-19 response. Regret bounds for a non-stochastic control setting [Hazan et al., 2020] with arbitrary bounded perturbations is a promising direction to explore. Additionally, we could investigate control strategies for applications such as online information propagation and regulation of occupational groups that exhibit Hamiltonian dynamics [Nutku, 1990].

## Data Availability

All data used for our experiments are publicly available.
Data sources include,
(i)Public Health England COVID-19 data
(ii)Covid19india data
(iii)The New York Times COVID-19 data

https://data.london.gov.uk/dataset/coronavirus--covid-19--cases

https://www.covid19india.org/

https://github.com/nytimes/covid-19-data

## Appendix

### A Extensions

#### Delayed SIR &SEIR Models

Certain infectious diseases have a significant incubation period when the individuals are infected but are not spreading the disease (non-infectious). The SEIR model [Hethcote, 2000] includes an additional **E** (exposed) compartment to model the incubation phase. This model is known to closely mimic the behaviour of the delayed SIR model [Kaddar et al., 2011]. When *β* follows Eqn. 2, the delayed SIR model readily maps to a delayed LV system with a non-preying growth period for the predators, which is a special case of the well-studied Wangersky–Cunningham systems [Martin and Ruan, 2001, Wangersky and Cunningham, 1957]. It can be shown that the modified delayed SIR system with a delay *τ* has the same equilibrium (*J*\*, *I*\*) = (*γN, r/e*), exhibits (unbounded) oscillations and permits control of the form Eqn. 3, where

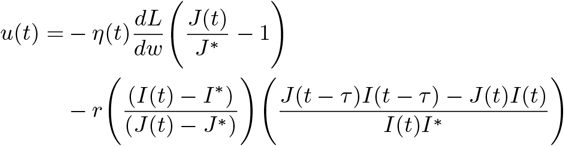

with *η*(*t*) > 0. There is a need for special handling when *J* approaches *J*\* with the behavior depending on *τ*.

#### Testing &Isolation Policy

Testing, tracing and isolation also play a critical role in regulating the epidemic. In terms of SIR and SEIR dynamics, the net effect of aggressive testing is minimizing the infectious period [Bar-On et al., 2020, Larremore et al., 2020]. This is analogous to the culling of predators (infectious population) by increasing the death rate for which there already exists multiple control mechanisms [Meza et al., 2005]. In particular, choosing *V* (**z**) = *L*(*w*(*J, I*)) as the CLF of interest, we obtain the control, 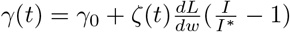, with *ζ*(*t*) > 0.

#### Non-stochastic Control

Figure 4 depicts the epidemic system from a control perspective. Theoretical guarantees of optimal control based on control Lyapunov functions rely on the assumptions of (a) deterministic transition dynamics, (b) stochastic perturbations *p*(*t*), and (c) exact control. In practice, however, the parameters of the SIR model such as the inverse infectious period (*γ*) vary with time, the perturbations due to in/out flow might be arbitrary, and the control is approximate because of public communication and compliance issues. To address these constraints, we need to continuously re-estimate the model parameters as well as the mapping between the restriction levels and *β* from historical data. In this online learning setting, the restriction levels can be viewed as bandit arms each associated with a specific transmission rate (or probability distribution) and compartmental populations correspond to the state. The arm-specific transmission rates are estimated in each step using observations based on an appropriate loss function. When the loss function is defined in terms of the Lotka-Volterra energy over a time horizon, this formulation can be viewed as a non-linear contextual bandit [Beygelzimer et al., 2011] and the properties of the LV energy permit derivation of regret bounds similar to the non-stochastic control framework for linear dynamical systems [Hazan et al., 2020].

**Figure 4:**
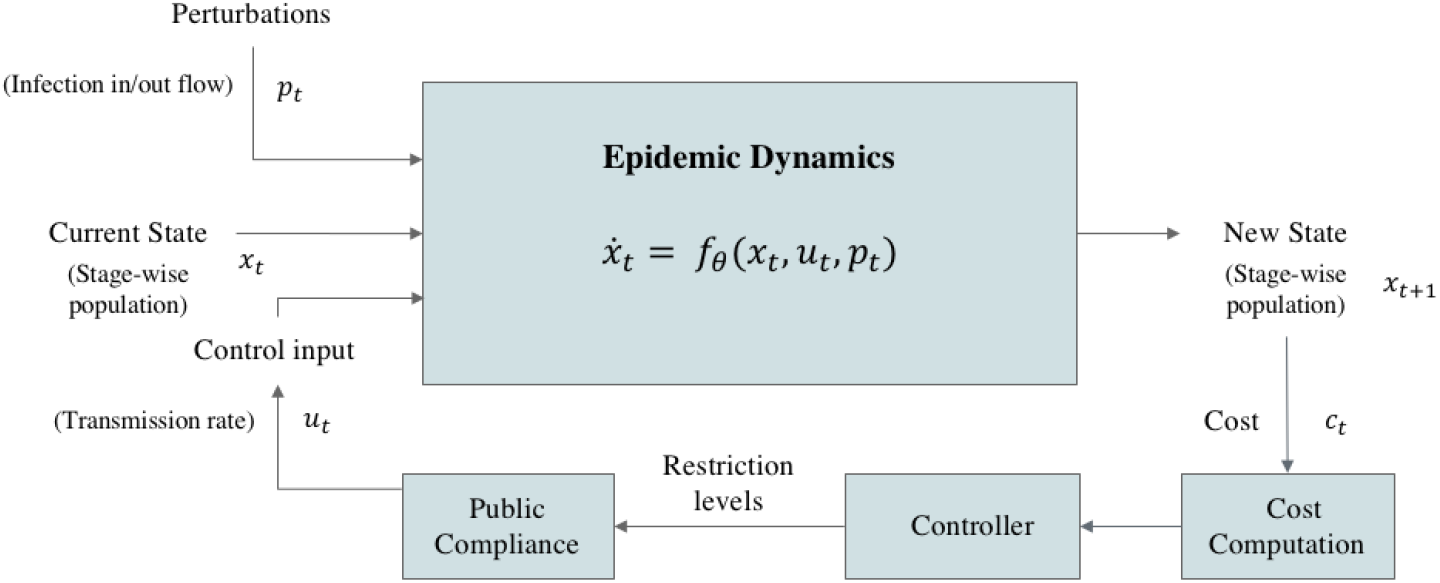
The control theory perspective of the epidemic dynamics.

### B Proofs

#### Definition 2

*[Boyce et al*., *2017] Let* **z**\* ∈ ℝ^*n*^ *be a critical point of a system of ODEs. The critical point* **z**\* *is stable if, for any ϵ* > 0 ∃ *δ* > 0 *such that if* **z** = *ϕ*(*t*) *satisfies* ||*ϕ*(0) − **z**\*|| < *δ then* ||*ϕ*(*t*) − **z**\*|| < *ϵ*, ∀ *t* > 0.

#### Proof of Theorem 1.1

At equilibrium (*J*\*, *I*\*), we have 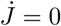 and *İ* = 0. From Eqn. 1, it follows that

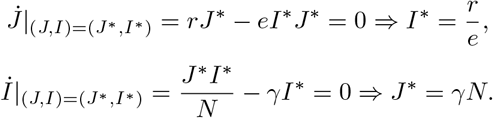

To prove the stability of the critical point at (*J*\*, *I*\*), let us consider the normalized variables *ϕ*(*t*) = (*x*(*t*), *y*(*t*)) where 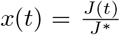 and 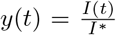. **z**\* = (1, 1) is the corresponding critical point.

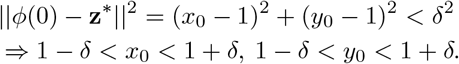

Let *f* (*s*) = *s* − log(*s*) − 1. Since *f* (*s*) is a convex function, 1 − *δ* < *s* < 1 + *δ*, implies *f* (*s*) < max {*f* (1 + *δ*), *f* (1 − *δ*)}. Denoting this bound by *D*_*max*_ implies *f* (*x*_0_) < *D*_*max*_ and *f* (*y*_0_) < *D*_*max*_

From Theorem 2.1, we note that

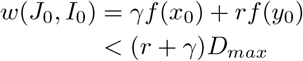

Denoting *w*_*b*_ = (*r* + *γ*)*D*_*max*_, from Theorem 2(a), we note that

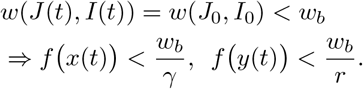

Given the nature of *f* (*s*), *f* (*s*) < *c* ⇒ *s*_*min*_ < *s* < *s*_*max*_ where (*s*_*min*_, *s*_*max*_) are the finite-valued roots of *f* (*s*) = *c*. Hence *x*(*t*) and *y*(*t*) are both bounded on either side. Consequently, (*x*(*t*) − 1, *y*(*t*) − 1) is confined to a bounded rectangle and thus,

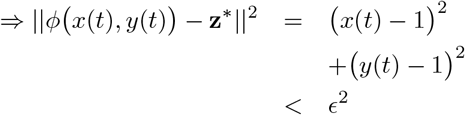

where *ϵ* can be directly expressed in terms of *δ* and vice versa.

Hence, from Definition 2, **z**\* = (1, 1) (or equivalently (*J*\*, *I*\*)) is a stable equilibrium.

#### Proof of Theorem 1.2

When initial state (*J*_0_, *I*_0_) is at equilibrium (*J*\*, *I*\*) = (*γN, r/e*), we have (*J* (*t*), *I*(*t*)) = (*γN, r/e*), ∀*t*. Hence,

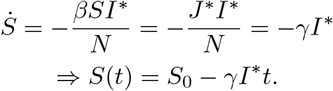

Similarly,

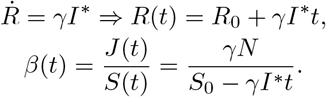

At *t* = *T*_*end*_, the susceptible population *S*(*t*) = 0. Hence,

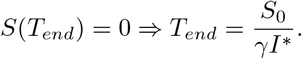

#### Proof of Theorem 2.1

The energy function of the LVSIR system in Figure 2(c) corresponds to

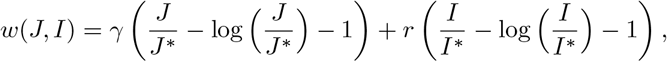

and the dynamics of *I, J* are given by

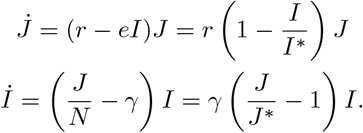

Considering the time derivative of *w*(*J, I*), we have

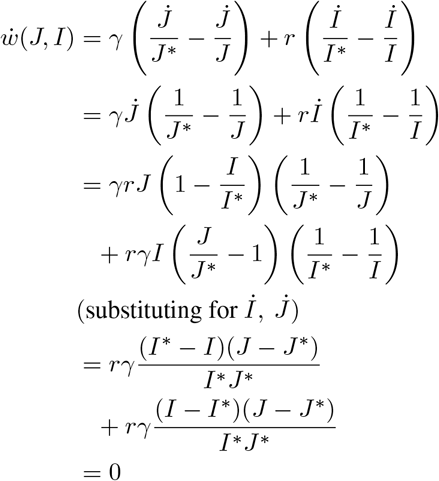

Hence, *w*(*J, I*) remains invariant throughout and is equal to *w*(*J*_0_, *I*_0_) = *w*_0_.

#### Proof of Theorem 2.2

Let *w*_0_ = *w*(*J*_0_, *I*_0_) be the energy associated with the modified SIR system in Figure 2(c). The conservation law implies that every valid state (*J, I*) corresponds to a point on the level curve given by

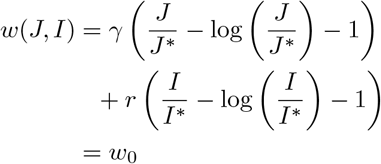

If *J, I* functions are continuous^6^, then these would be periodic functions. In terms of normalized variables, 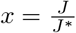 and 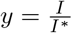, the phase plot reduces to

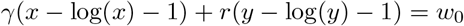

Consider the continuously differentiable function *f* (*z*) = *z* − log(*z*) − 1 defined on ℝ_++_. Since 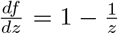 and 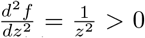, *f* (*z*) is a convex function with a single global minimum at *z* = 1 corresponding to *f* (1) = 0. Hence *f* (*z*) ≤ *c* ⇒ *z*_*min*_ ≤ *z* ≤ *z*_*max*_ where (*z*_*min*_, *z*_*max*_) correspond to the roots of *f* (*z*) = *c*.

To identify the extreme *x* values, we observe that

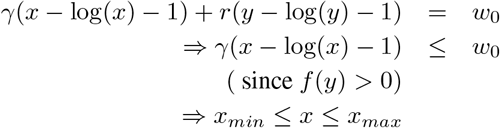

where (*x*_*min*_, *x*_*max*_) are roots of 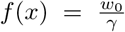. Both the extreme values of *x* are realized for *y* = 1. Similarly, the extreme values of *y* are attained for *x* = 1 and given by (*y*_*min*_, *y*_*max*_) which correspond to the roots of 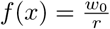.

#### Proof of Theorem 2.3

The period of a Lotka-Volterra system has been derived in multiple works Shih [1997]. We include the below proof based on Hsu’s method Hsu [1983] for completeness.

Let 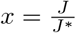 and 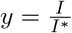. Then we have

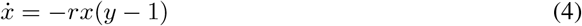

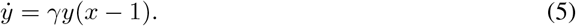

From 4, we have

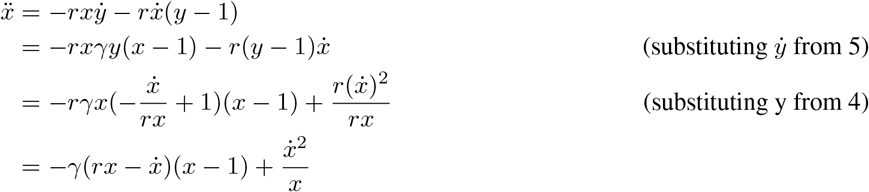

Thus,

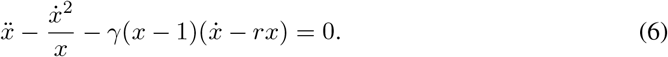

Let *z* = log(*x*). Then, 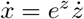 and 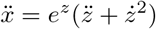.

From 6, we have

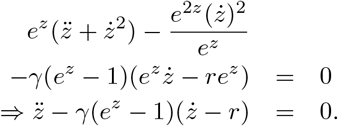

Choosing 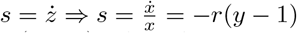.

Let *w*_0_ = *w*(*J*_0_, *I*_0_). Then, the trajectory corresponds to

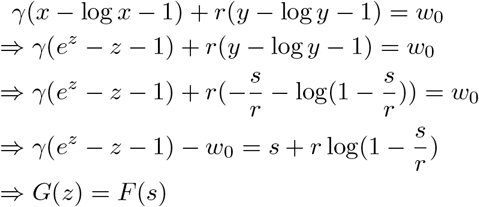

where *G*(*z*) = *γ*(*e*^*z*^ − *z* − 1) − *w*_0_ and 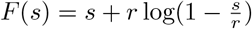.

Let *F*_1_(*s*), *F*_2_(*s*) be the restrictions of *F* (*s*) for the lower and upper parts of the phase plot. Then the time period for the lower section is given by

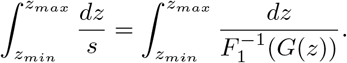

The total time for both the lower and upper section is given by

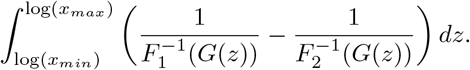

When *w*_0_ ≃ 0, linearization is possible. Simplifying the trajectory *F* (*s*) = *G*(*z*) using the approximations 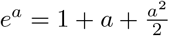 and 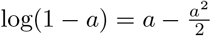, we have

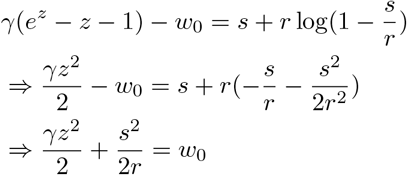

Essentially, we have an elliptical curve with *x, y* following sinusoidal behavior with a period 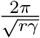.

#### Proof of Theorem 2.4

Assuming a continuous form for *J*, we observe that

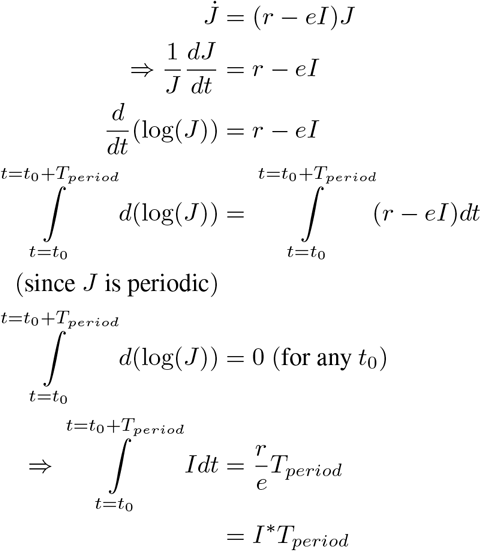

In other words, *I*\* is also the average value of *I* in each cycle.

Considering the time derivatives of *S* and *I*, we have 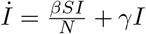 and 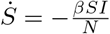.

Let Δ*S* be the drop in *S* during a single cycle starting at any *t*_0_, then

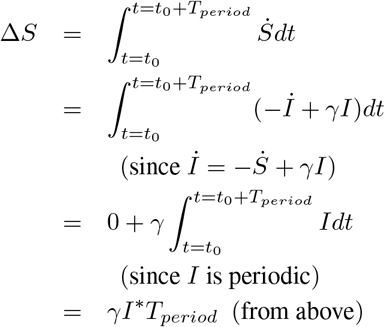

#### Proof of Theorem 4.

Assuming a proportional additive control on *β* of the form 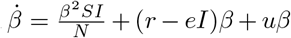, the variation of the susceptible contacts *J* is given by

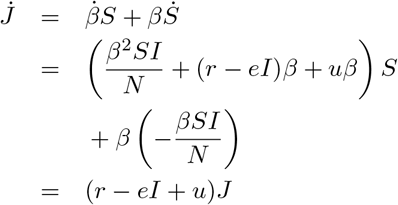

Let **z** = (*J/J*\* − 1, *I/I*\* − 1) = (*z*_1_, *z*_2_) so that **z** = (0, 0) corresponds to the equilibrium state. Then *w*(*J, I*) = *γ*(*z*_1_ − log(1 + *z*_1_)) + *r*(*z*_2_ − log(1 + *z*_2_)). For *V* (**z**) = *L*(*w*(*J, I*)) to be a control-Lyapunov function, we require 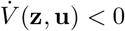.

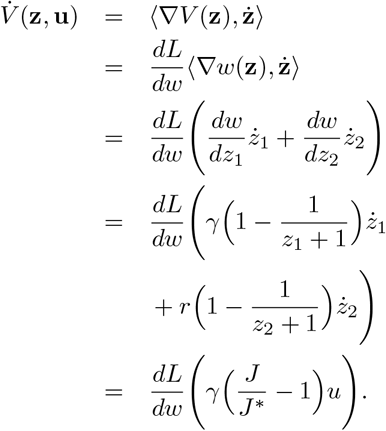

When the control is chosen as

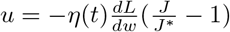 and *η*(*t*) > 0, ∀*t*,

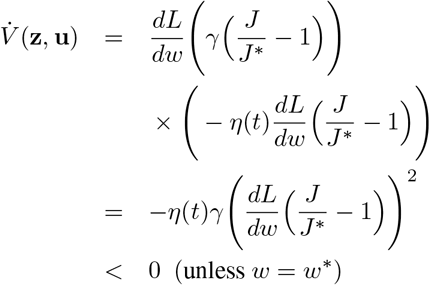

Hence, Artstein’s theorem guarantees convergence to the equilibrium.

### C Parameter Choices

Table 4 enumerates the detailed parameter choices and the relevant assumptions for the different regions that can be broadly divided into three groups. We now discuss the rationale for the various parameter choices.

**Table 4:** State of the epidemic, public health requirements, and COSIR parameters used for the simulations in Section 7.

| Factor | Notation | Hypothetical | India | Netherlands |
| --- | --- | --- | --- | --- |
| Population | $N$ | 13M | 1.366B | 17.1M |
| Stage of pandemic | $(S_{curr}, I_{curr})$ | (12.6M, 0.2M) | (1.36B, 279k) | (17.09M, 6.7k) |
| Bed Occupancy level (Phillip et al. [1984]) | $\kappa$ | - | 0.76 | 0.76 |
| Medical Capacity (per capita) (capacity [2020]) | $p$ | - | $5.3 \times 10^{-4}$ | $3.32 \times 10^{-3}$ |
| Hospitalization ratio | $h$ | - | 2% | 6.6% |
| Time spent in hospital | $T_{hosp}$ | - | 20 days | 20 days |
| Target manageable infections | $I_{avg}^{target}$ | 0.15M | 6.88M | 0.16M |
| Target max to average ratio | $I_{max}^{target} / I_{avg}^{target}$ | 1.3 | 1.13 | 1.13 |
| Infection-Fatality ratio (Meyerowitz-Katz and Merone [2020], Mukhopadhyay and Chakraborty [2020], Murhekar et al. [2020]) | IFR | - | 0.002<sup>7</sup> | 0.0068 |
| Death detection ratio | DDR | - | 0.3 | 1 |
| Reported Infections on 1st Dec 2020 (Worldometer [2021], COVID19India [2020]) |  | - | 9.5M | 527k |
| Reported Fatalities on 15th Dec 2020 (Worldometer [2021], COVID19India [2020]) |  | - | 144k | 10k |
| Under reporting factor | URF | - | 25.28 | 2.83 |
| Periodicity | $T_{period}$ | 7 days | 7 days | 7 days |
| Reciprocal of infectious period | $\gamma$ | 0.2 | 0.2 | 0.2 |
| LV Reproduction rate | $r$ | 4.2 | 4.2 | 4.2 |
| Consumption rate | $e$ | $2.8 \times 10^{-5}$ | $6.1 \times 10^{-7}$ | $2.6 \times 10^{-5}$ |
| Initial transmission rate | $\beta_0$ | 0.2 | 0.2 | 0.2 |
| Learning rate | $\eta$ | 5 | 5 | 5 |

a. **Target Hospitalization &Infectious levels** There are multiple ways to arrive at an acceptable level for infectious population 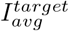 based on socioeconomic impact considerations [BMGF, 2020]. When the focus is on medical capacity constraints, this target level can be set chosen so that the available medical capacity can meet the hospitalization requirements at a steady state. Let *p* denote the per-capita hospital bed capacity of a region with population *N, κ* the bed occupancy level for normal functioning, and *T*_*hosp*_ the average duration of hospitalization. Then, a manageable hospitalization inflow that will not overwhelm the medical infrastructure is given by 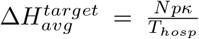. As described earlier in Section 7, assuming SIR dynamics, the new hospitalizations would be given by *γhI*. Hence, we choose the target infectious level as 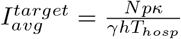.To concretely instantiate these choices for various regions, we use the populations of the regions *N*, the per-capita medical capacity capacity [2020] assuming an average hospitalization period *T*_*hosp*_ = 20 days and hospitalization ratio between 2 − 6.6% based on literature. Note that these parameters are likely to vary across regions and there is a significant uncertainty in the estimates across multiple studies. However, the key insights on the relative behavior of the transmission control policies hold true regardless of the specific choices of the parameter values.
b. **Under Reporting Factor** While comparing real observed case counts with simulations, a critical factor to consider is the level of under reporting. This factor was computed based on the assumption that the infection fatality rate (*IFR*) and death detection rate (*DDR*) are largely variant within a region and there is a steady lag of 2 weeks between infection and the associated fatalities. In particular, we estimate the under reporting factor (URF) as

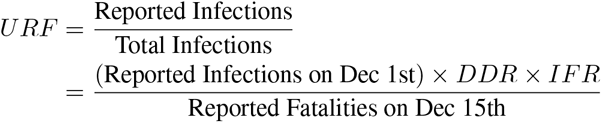 The purpose of computing URF is to figure out the appropriate scaling of real cases for a fair comparison with the simulated results.
c. **SIR &CoSIR Parameters** The primary parameter of interest in the SIR model apart from the transmission rate is *γ*, i.e., reciprocal of the infectious period, which was chosen to be 0.2 across all simulations since it is a disease-specific factor. For practical implementation of a control policy, a periodicity aligned with typical economic activity is preferable and hence, we choose a periodicity of *T*_*period*_ = 7 days for the CoSIR model. Assuming the maximum value of infections to be 13% higher than the average and following Algorithm 1, we obtain the LV reproductive rate *r* = 4.2. We choose the steady state infectious level to be the target infectious level, i.e., 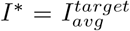 and the consumption rate is given by *e* = *r/I*\*. To ensure effective control, we pick an aggressive learning rate *η* = 5.

### D Additional Experiments

To assess the impact of the different transmission control policies on socioeconomic activity, we consider the variation of the restriction level, i.e., *β* over time. We also compute for each policy, the total number of days where the transmission rate associated with the policy was higher (i.e., less restrictive) than a particular *β* and discuss the associated implications on socioeconomic impact.

Figures 5(a-b) show the variation of the transmission rate (*β*) with time for four intervention policies (`CoSIR`, `CoSIR-approx`, `PL-high`, `PL-low`). All the approaches involve alternating between different levels of restrictions but the CoSIR based approaches adapt these levels as the susceptible population decreases over time. In the case of Netherlands, this approach nearly permits a return to no restrictions by April 2021 even without other interventions such as vaccination (conditioned on the assumptions on the acceptable hospitalization levels).

**Figure 5:**
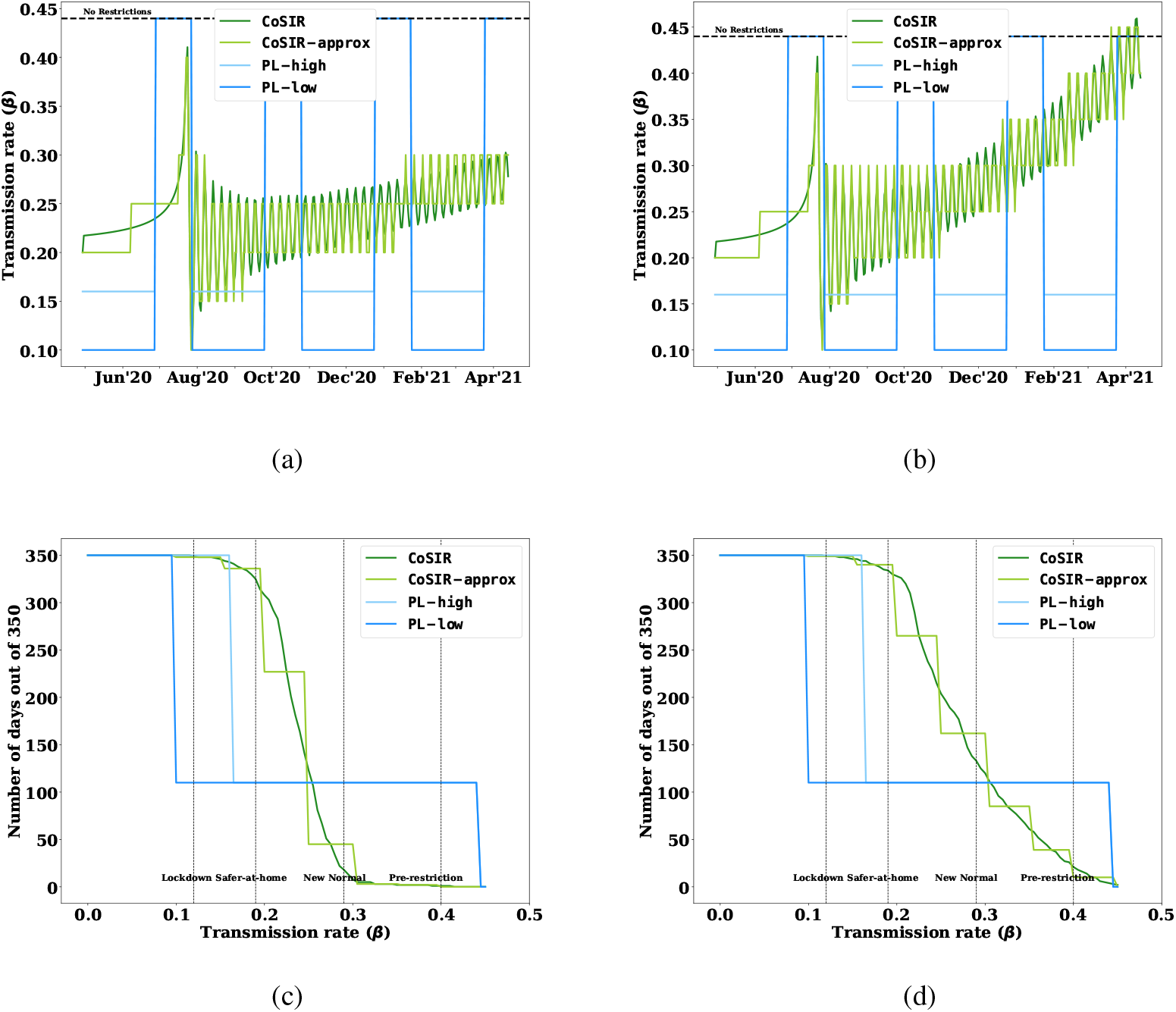
Plots (a-b) depict the variation of policy transmission rate with time. Plots (c-d) show the restrictiveness profile (number of days where the policy is less restrictive than a specified *β*) for the periodic lockdown and CoSIR-based policies for India and Netherlands

Figures 5(c-d) show the number of days where a control policy allows a transmission rate higher than a particular *β*, i.e., in other words allows all socioeconomic activities that necessitate a transmission rate of *β*. The periodic lockdown policies tend to have a discontinuous profile while the CoSIR based policies allow a more continuous transition. In particular, a large number of vital socioeconomic activities correspond to *β* between “safer-at-home” and “new normal”. The CoSIR based policies clearly dominate in this region. Given a socioeconomic model that maps *β* to socioeconomic costs and the distribution of the transmission rate *β*, we could quantitatively evaluate the socioeconomic impact of the various policies. We do not provide such an impact analysis due to the complexity of socioeconomic modeling in the context of significant variations between high income and low-middle income regions [Zachary and Mobarak, 2020]. However, since small relaxations in restriction levels in the range between “stay/safer at home” policies and “new normal” do seem to allow a substantial increase in socioeconomic activity with diminishing returns beyond that stage, the differences in the *β* profile are likely to be amplified when the socioeconomic impact is considered. Note that if the simulation were to cover the entire duration of the pandemic, the CoSIR based approaches would turn out to be more relaxed than the fixed periodic lockdowns.

Figure 6 show the LV Energy curve with respect to time for the different policies. The energy, which denotes how far the systtem is from the target values, quickly comes down for the CoSIR variants. Table 5 shows the total energy of each of the policy over the entire duration of 350 days. The total energy of the CoSIR variants is significantly low which signifies that the system remained close to the target levels.

**Figure 6:**
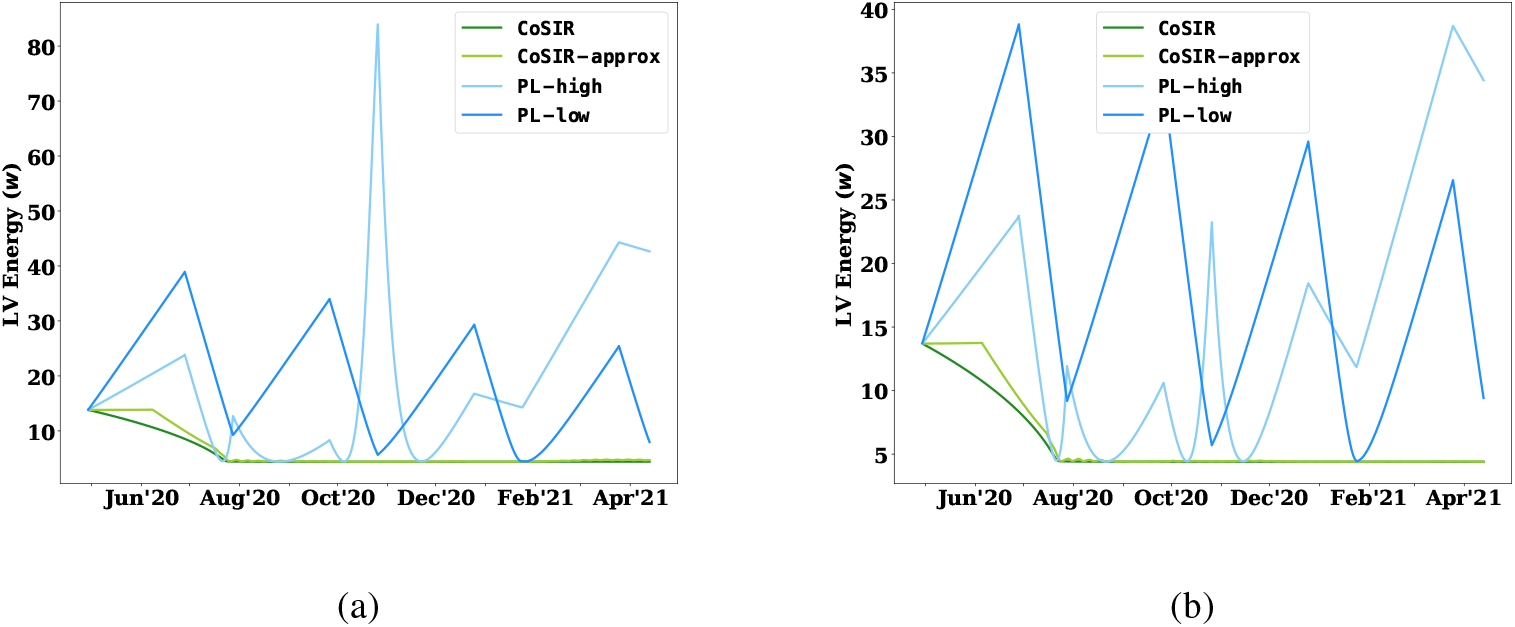
Plots (a-b) depict the variation of LV Energy with time corresponding to all policies for India and Netherlands respectively.

**Table 5:** Total LV Energy for the entire duration of 350 days corresponding to all policies for India and Netherlands respectively.

| Policy | India | Netherlands |
| --- | --- | --- |
| CoSIR-ideal | 2026.6 | 2009.7 |
| CoSIR-approx | 2184.6 | 2133.2 |
| PL-high | 6561.9 | 5416.7 |
| PL-low | 6744.9 | 6826.6 |

#### D.1 Empirical results till April 2021

Given the heavy interest in the subsequent waves of infections in India and Europe, in the interest of transparency, we also have included plots showing the real outcome till April (Figure 7). However, as per the discussion in Section 9, one needs to be mindful that the proposed control policy (CoSIR) applies only when SIR dynamics are valid. In the first phase of the pandemic (most of 2020) when reinfection and vaccination rate was negligible, this is largely true but it not the case for the most recent phase. The right approach is to consider a new compartmental model that incorporates the reinfection and vaccination dynamics and derive the corresponding optimal control, which is a direction that we are exploring.

**Figure 7:**
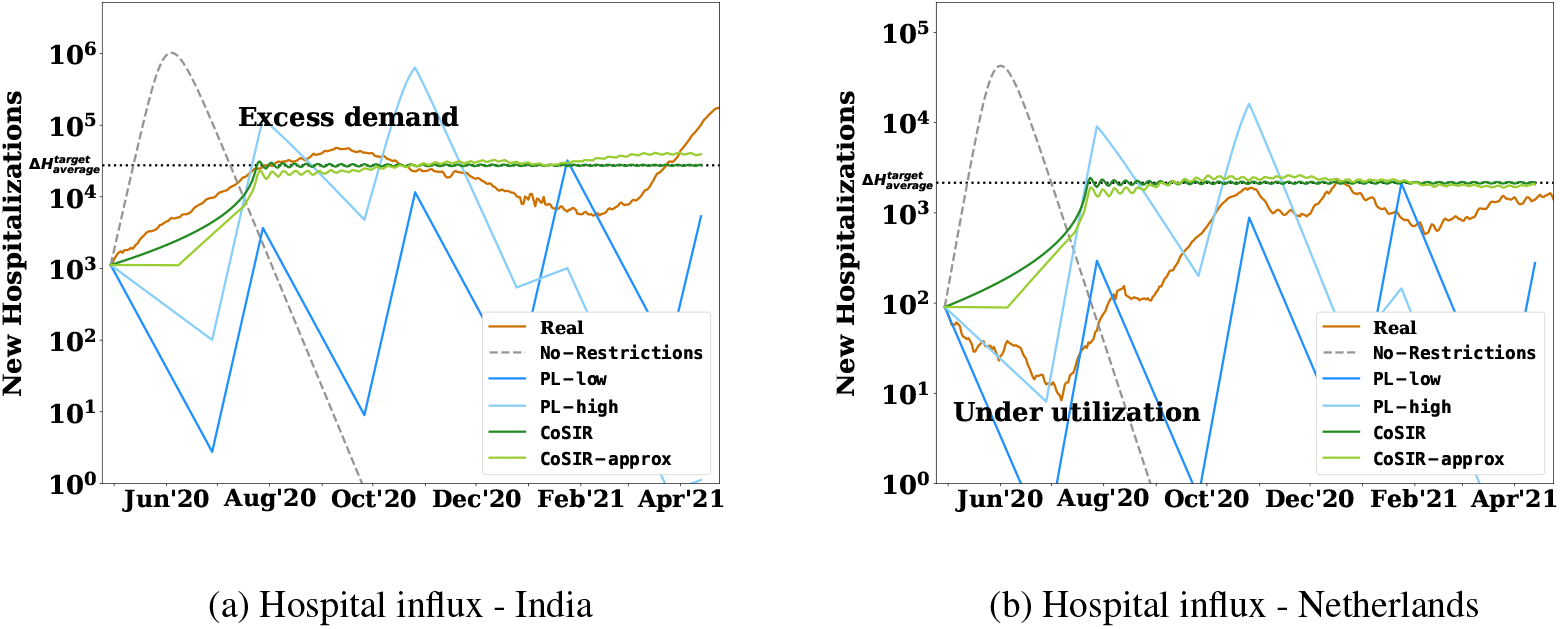
New hospitalization influx with various transmission control policies for two regions: India-IN and Netherlands-NL with real data till April 2021

### E Societal Impact

Epidemic and population control have applications in multiple domains such as public health, public communications, viral marketing, regulation of economic activity in multi-sided market places, all of which have an immediate and significant societal impact. Better understanding and controllability of these macro-systems that impact human lives definitely holds the promise of positive outcomes. However, as in the case of generic AI techniques, the proposed control methods create an empowered artificial agency that does entail multiple risks for the society including but not limited to:

- Inappropriate and unsafe use of control techniques due to poor understanding of the limitations and underlying assumptions of the proposed methods (see Section 9) as well as inadequate safeguards to address these constraints, especially in areas related to goal/input specification, data observability, and model dynamics. Such misuse (e.g., an inappropriate target infectious level) can happen even with benevolent controllers and the best of intentions.
- Use of control techniques for objectives contrary to societal good such as propagating misinformation on social media, marketing harmful products, and bio-warfare.
- Increase automation of higher order decision that could lead to more efficiency on one end, but also loss of employment and further empowerment of capital relative to human resources.

Both the benefits and risks associated with the proposed techniques are amplified by the fact that the immediate applications such as control of pandemics and online discourse pertain to regulation of macro-systems affecting large human populations.

### F Miscellaneous

This section includes miscellaneous content and larger versions of figures for the readers benefit. Table 6 contains an example of restriction levels and the corresponding effective transmission rates (*β*).

**Table 6:** Example of restriction levels recommended by public health authorities [COVID-AMP, 2020] and the corresponding effective R value and transmission rate (*β*) for COVID-19.

| Restriction level | Effective R value | COVID-19 Transmission rate ($\beta$) | Description |
| --- | --- | --- | --- |
| Early event (Pre-restrictions) | 2.52 | 0.4 | No policy interventions. |
| Lockdown | 0.84 | 0.12 | Residents are either not allowed to leave their residence or may leave only for essential functions. |
| Stay-at-home | 0.95 | 0.14 | Stay-at-home order includes closure of schools and private sector, and restrictions on mass gatherings. |
| Safer-at-home | 1.26 | 0.19 | Relaxed stay-at-home order which includes closure of schools, but restrictions on mass gatherings and private sector may be relaxed. |
| New normal | 1.83 | 0.29 | Stay-at-home lifted or relaxed with some restrictions on mass gatherings and private sector. Schools may or may not be re-opened. Safeguards such as face coverings encouraged. |

**Figure 8:**
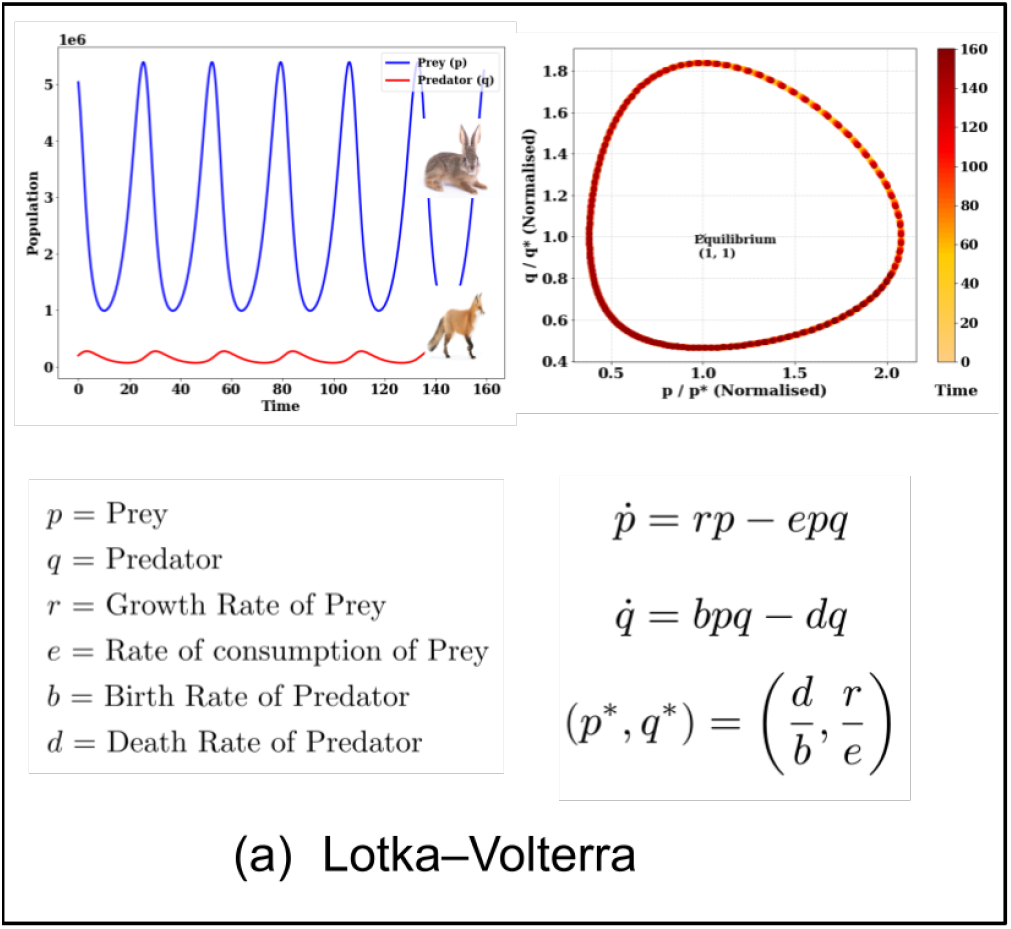
(a) Lotka-Volterra system and evolution of predator (fox) and prey (hare) population over time.

**Figure 9:**
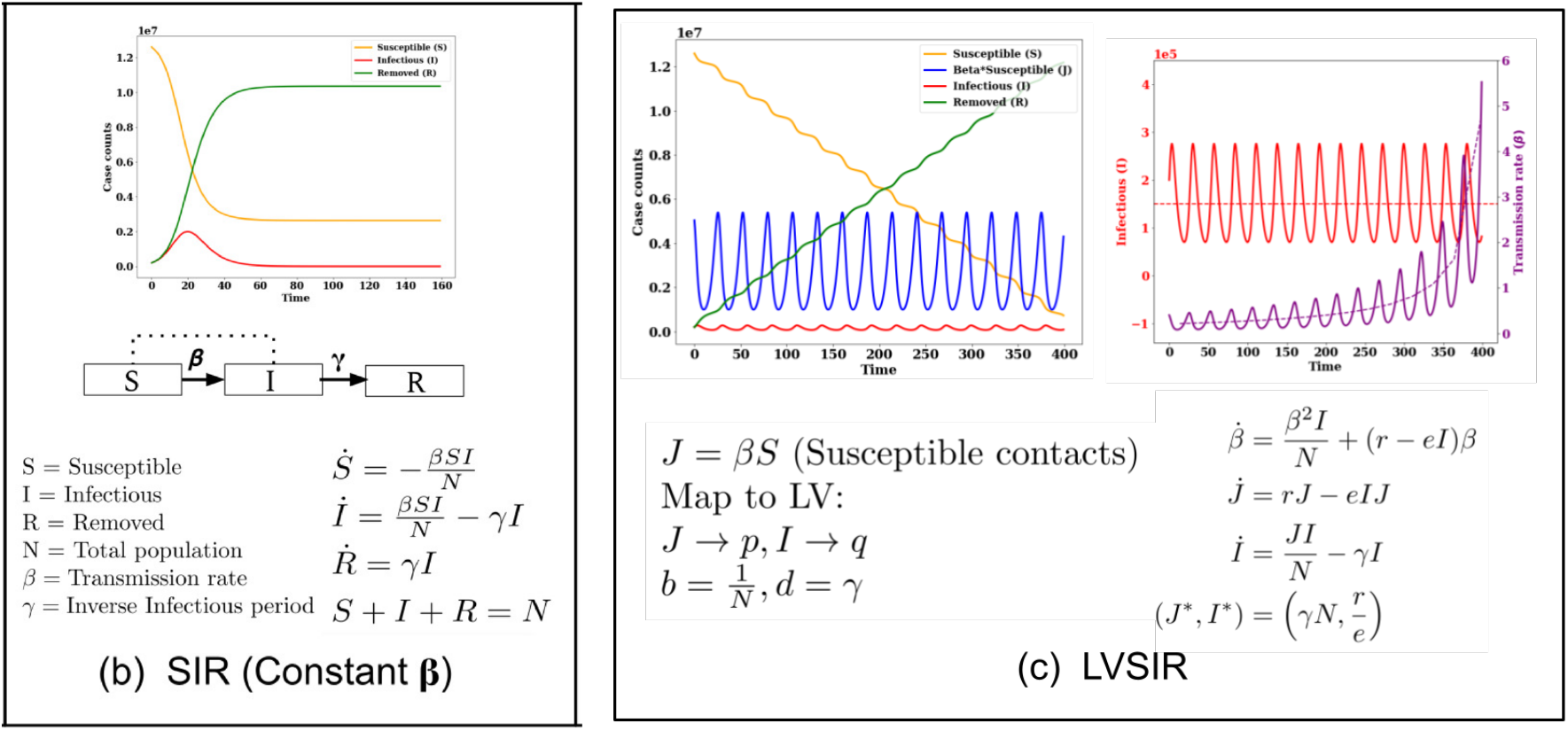
(b) SIR compartmental dynamics for epidemiological modeling. (c) Mapping of SIR model to LV system and the behaviour of case counts *S, J, I, R* and transmission rate *β*.

**Figure 10:**
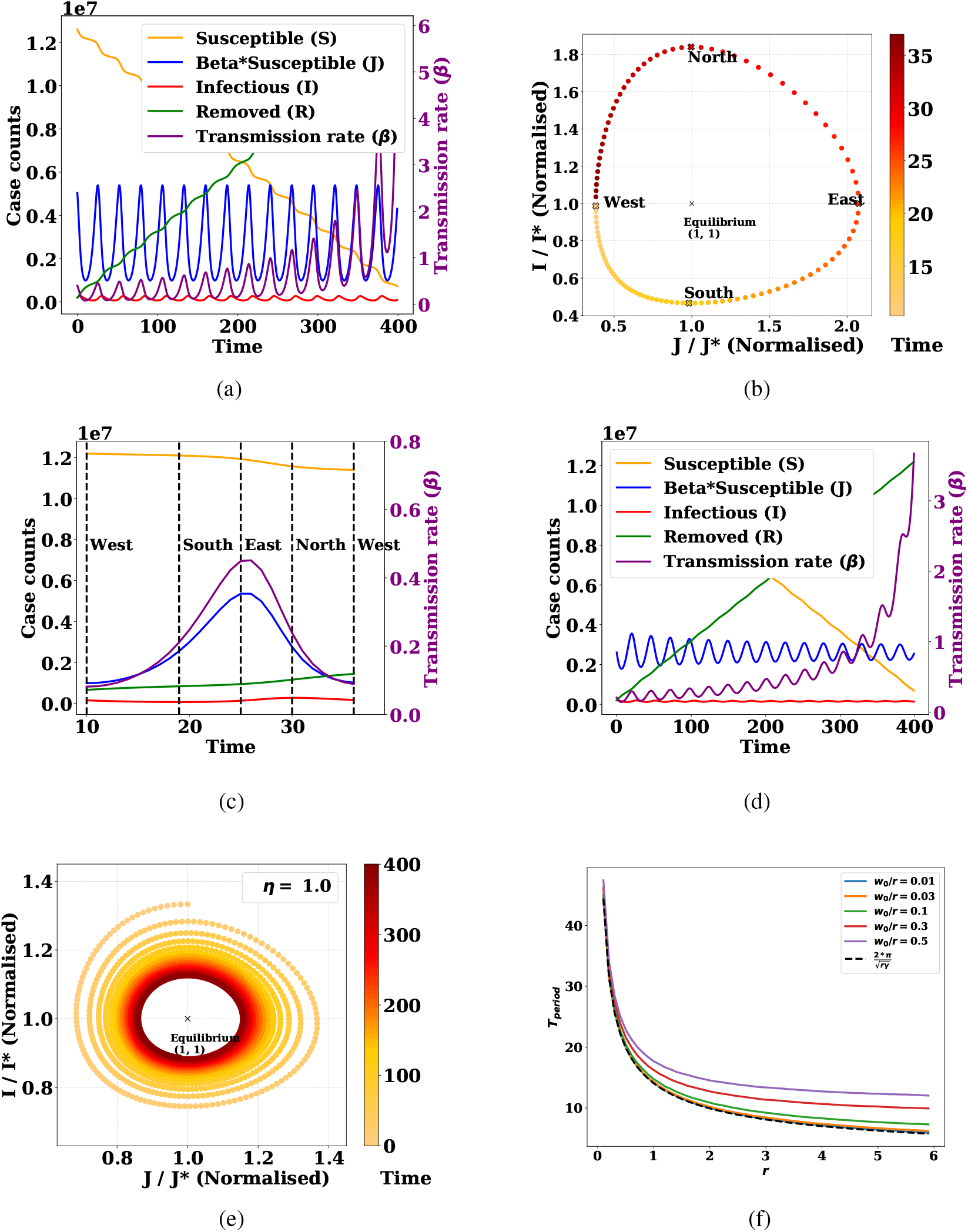
(a-c) System (*S, J, I, R, β*) evolution when *β* follows Eqn. 2 [LVSIR] with its phase plot and single cycle annotated with extreme points, (d-e) System evolution when *β* follows Eqn. 3 [CoSIR] and learning rate *η* = 1, (f) Dependence of *T*_*period*_ on *r* for different choices of *w*_0_*/r*.

**Figure 11:**
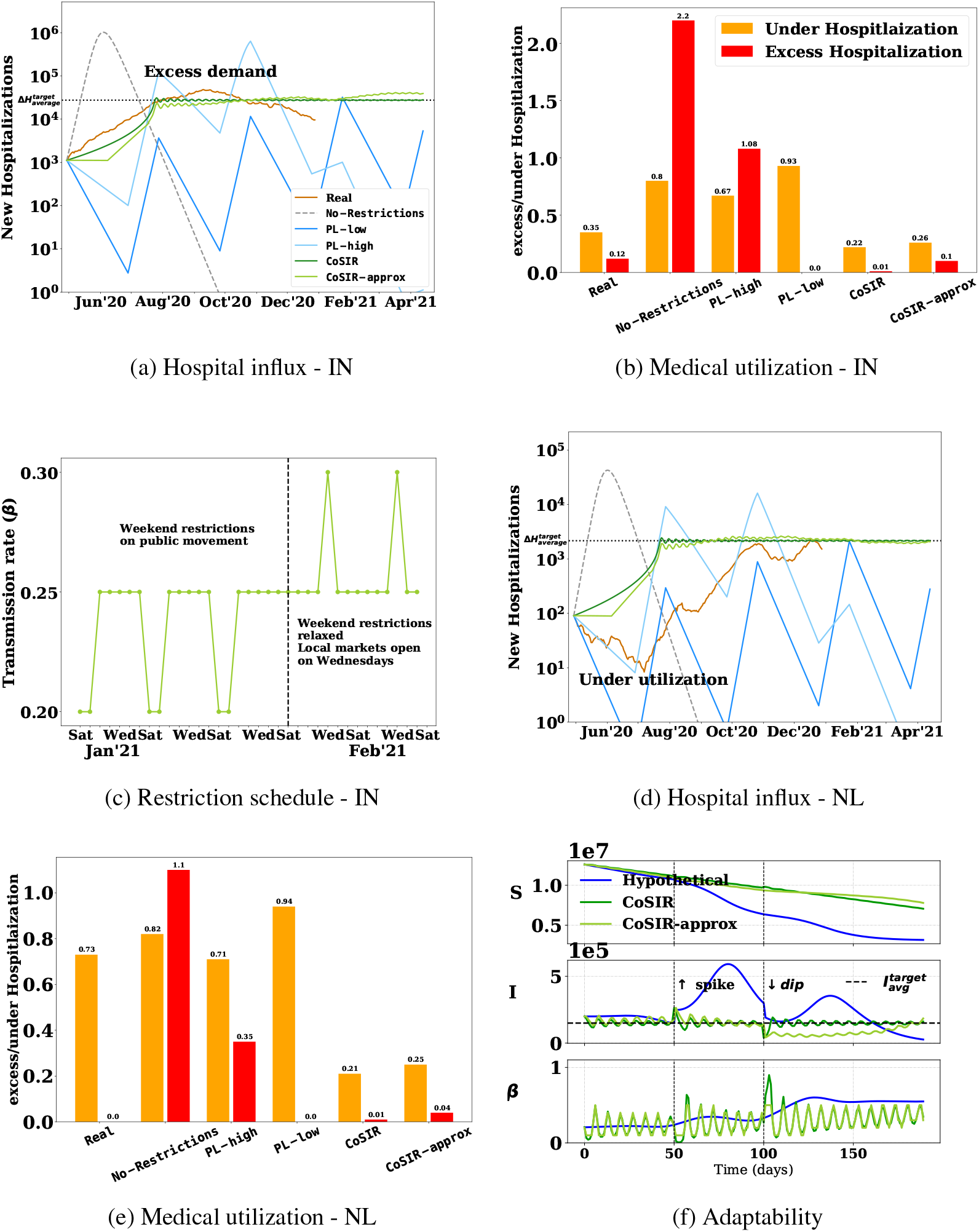
(a-b, d-e) New hospitalization influx and excess/under hospitalization with various transmission control policies for two regions: India-IN and Netherlands-NL, (c) `CoSIR-approx` schedule (IN) for a 5-week period (see Appendix F for full range) with a possible mapping to real restrictions, (f) Disease progression indicates that `CoSIR` variants adapt to target levels and also to a sudden increase (*t* = 50) or decrease (*t* = 100).

## Footnotes

2 Extensions to other models with an incubation period (e.g., SEIR) as well as control of other variables such as the infectious period are presented in Appendix A.

3 Mapping restrictions to transmission is the focus of multiple works [Lehner, 2020] and discussed in Sec 6.

4 Please see Appendix B and F for proofs and larger figures respectively. Webapp: http://cosir.herokuapp.com/

5 LV reproduction rate *r* is different from that of the SIR reproduction number.

6 Note that *J, I* are actually discrete population counts and not continuous functions.

7 Since these estimations vary, we choose an intermediate value.

